# Intermediate levels of asymptomatic transmission can lead to the highest levels of epidemic fatalities

**DOI:** 10.1101/2022.08.01.22278288

**Authors:** Sang Woo Park, Jonathan Dushoff, Bryan T. Grenfell, Joshua S. Weitz

## Abstract

Asymptomatic infections have hampered the ability to characterize and prevent the transmission of SARS-CoV-2 throughout the ongoing pandemic. Even though asymptomatic infections reduce severity at the individual level, they can make population-level outcomes worse if asymptomatic individuals—unaware they are infected—transmit more than symptomatic individuals. Using an epidemic model, we show that intermediate levels of asymptomatic infection lead to the highest levels of epidemic fatalities when the increase in asymptomatic transmission, due either to individual behavior or mitigation efforts, is strong. We generalize this result to include presymptomatic transmission, showing how intermediate levels of non-symptomatic transmission can lead to the highest levels of fatalities. Finally, we extend our framework to illustrate how the intersection of asymptomatic spread and immunity profiles determine epidemic trajectories, including population-level severity, of future variants.

---

SARS-CoV-2 has had devastating effects at the population level. However, many individuals experienced mild cases, making it harder to estimate the magnitude of spread and fatality rate [1]. The ratio of fatalities to documented cases (the case-fatality rate, CFR) is typically between 1%–4%, varying across population because of testing patterns, treatment practice, case definitions, and other factors [2, 3, 4]. But many infections are never documented; the ratio of fatalities to total infections (the infection fatality rate, IFR) has been estimated to be closer to 0.5%–1% for pre-vaccinated populations whose demographics are similar to those of the United States [5]. This means that more than 99% of individuals infected with COVID-19 will survive. Moreover, at least half of the infections are sufficiently mild that they could be classified as subclinical or even asymptomatic.

Early in the pandemic, a COVID-19 outbreak on the Diamond Princess cruise ship played a critical role in understanding the role of asymptomatic infections in the spread of SARS-CoV-2; the outbreak occurred among 3711 passengers and crew, of whom 634 individuals tested positive by 20 February 2020 [9]. It has been estimated that 75% (95% C: 70%-78%) of all infections on the cruise ship were asymptomatic (Fig. 1A) with about half of total infections undetected [6]. The relative transmission rate of asymptomatic individuals aboard the Diamond Princess was not well constrained, but low relative transmission rate (below 25%) by asymptomatic individuals was ruled out because it required unrealistically high transmissibility for symptomatic individuals (Fig. 1B). Modeling studies have typically assumed that transmissibility is lower for asymptomatic than for symptomatic individuals; assumptions have ranged from 10%–100% [10, 11]. Similarities in viral load trajectories of asymptomatic and symptomatic individuals provide indirect support for the transmissibility of asymptomatic individuals (Fig. 1C, [7]); however, differences between inferred total viral load from Ct values and infectious viral load add uncertainties to how well asymptomatic individuals can transmit relative to that of symptomatic individuals [12]. We note also that asymptomaticity is expected to be more heterogeneous in a diversity of outbreak settings [13]. For example, during the early pandemic, Davies *et al*.’s analyses of surveillance data across six countries revealed that older individuals are less likely to have subclinical infections (Fig. 1D), providing indirect evidence for heterogeneity in asymptomaticity [8]. Differences in contact rates between age classes further contribute to the heterogeneity in asymptomatic transmissibility. For now, we primarily focus on a homogeneous population and return to the age effect in discussing our model-based findings.

**Figure 1:**
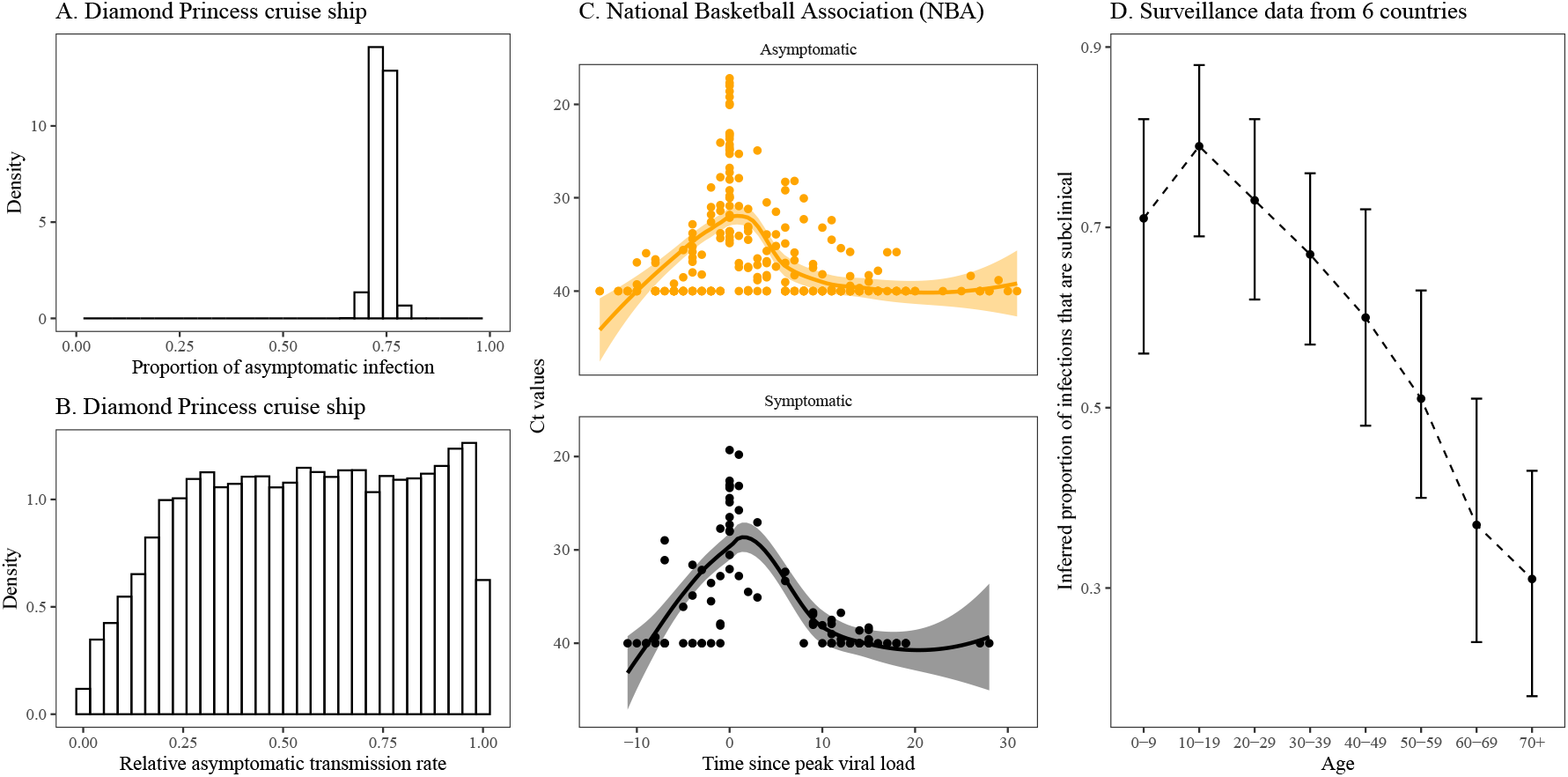
Asymptomatic transmissibility of SARS-CoV-2. (A) Posterior estimates of the proportion of asymptomatic infections from the Diamond Princess cruise ship [6]. (B) Posterior estimates of the ratio *θ*_*a*_ of the transmission rates between asymptomatic and symptomatic individuals from the Diamond Princess Cruise Ship [6]. Bar charts represent the posterior distributions. Symptomatic individuals were assumed to transmit at rate *β*(*t*) for an average of 2.9 days, followed by a pre-symptomatic stage with an average of 2.1 days. Asymptomatic individuals were assumed to transmit at rate *θ*_*a*_*β*(*t*) for an average of 5 days. Both estimates are publicly available with further details in [6]. (C) Viral load trajectory data from players, staff, and vendors of the National Basketball Association (NBA). Points represent each Ct measurement. Lines and shaded areas represent the smooth trajectories estimated via LOESS and the associated 95% confidence intervals. Data are publicly available in [7]. (D) Inferred proportion of infections that are subclinical for each age group using surveillance data from six countries (China, Italy, Japan, Singapore, South Korea, and Canada) [8].

Despite quantitative uncertainties in asymptomatic transmissibility, individuals infected asymptomatically with SARS-CoV-2 can still transmit to others. This means that the presence of asymptomatic infections may have countervailing effects at the population level. On one hand, an asymptomatic infection means that the individual infected avoids hospitalization and death. On the other hand, asymptomatic infections are less likely to be detected [14], meaning that asymptomatic individuals are less likely to take precautions and relatively more likely to infect others; asymptomatic SARS-CoV-2 infections present additional challenges to managing overall disease burden due to the possibility of long COVID [15]. Altogether, the prevalence of asymptomatic infections could paradoxically make population-level outcomes worse than if SARS-CoV-2 was more dangerous at the individual level.

To explore this idea, we propose a simple epidemic model, in which infected individuals can be asymptomatic or symptomatic, with probabilities *p* and 1 − *p*, respectively (Fig. 2A). Asymptomatic individuals always recover, whereas a fraction *f* of symptomatic individuals die. Asymptomatic and symptomatic individuals can also have different infection characteristics, including their transmission rates (*β*_*a*_ and *β*_*s*_) and removal rates (*γ*_*a*_ and *γ*_*s*_). Our key assumption is that symptomatic individuals take greater precautions than do asymptomatic individuals (e.g., via reducing contacts or increased mask-wearing) and therefore reduce their transmission rate by a fraction *δ*; the parameter *δ* may also capture intervention measures that target symptomatic individuals, such as symptom-based isolation. We note that intervention measures that target asymptomatic infections would reduce the effective value of *δ*—for example, frequent testing and isolation may effectively increase the removal rate *γ*_*a*_ of asymptomatic individuals. For our main simulations, we assume that asymptomatic individuals have a lower reproduction number—this is modeled by assuming lower transmission rates for asymptomatic individuals (*β*_*a*_ = 0.75*β*_*s*_) and equal removal rates (*γ*_*a*_ = *γ*_*s*_). We evaluate the effects on population-level mortality of changing the asymptomatic proportion *p* while holding the fatality rate for *symptomatic* cases, *f*, constant (the IFR (1 − *p*)*f* thus decreases as *p* increases).

**Figure 2:**
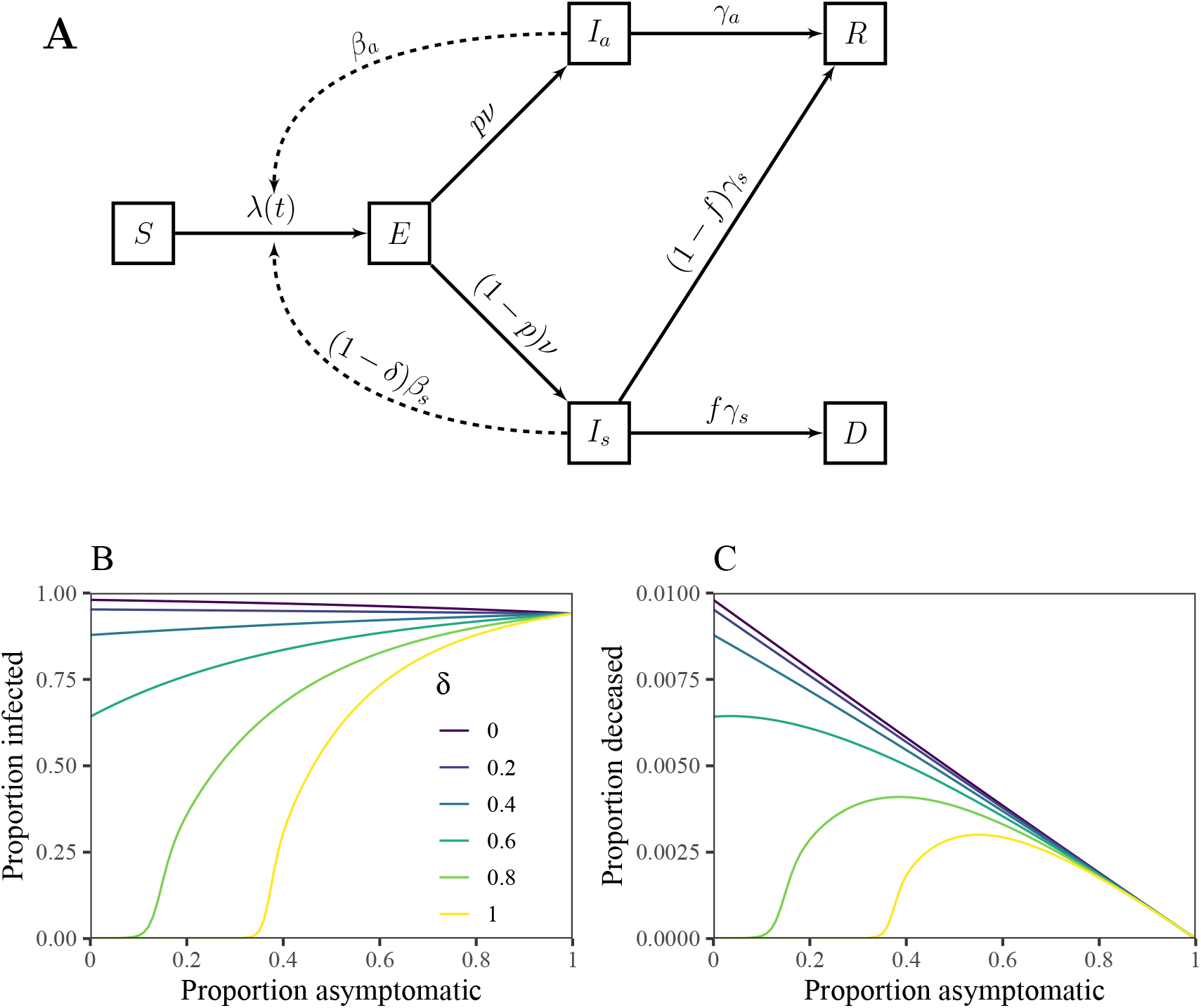
Schematic diagram and simulations of a model with asymptomatic transmission and symptom-responsive transmission reduction. (A) *S* represents susceptible individuals; *E* represents exposed individuals; *I*_*a*_ represents asymptomatic individuals; *I*_*s*_ represents symptomatic individuals; *R* represents recovered individuals; and *D* represents deceased individuals. See Methods for model details. (B) Total infections as a function of the proportion of asymptomatic infections *p* across a wide range scenarios for *δ*. (C) Total deaths as a function of the proportion of asymptomatic infections *p* across a wide range scenarios for *δ*. We simulate the model for 365 days, assuming *β*_*s*_ = 0.8*/*day, *β*_*a*_ = 0.75*β*_*s*_, *ν* = 0.5*/*day, *γ*_*s*_ = *γ*_*a*_ = 0.2*/*day, and *f* = 0.01, and an initial exposed proportion of 10^−4^. See Supplementary Text for model details and Supplementary Table S1 for parameter descriptions and values.

Fig. 2B–C shows simulated epidemic outcomes using parameters similar to those of the originating strain of SARS-CoV-2, without any mitigation other than that individuals who are symptomatic reduce their transmission rate by *δ*. For this model, the basic reproduction number is given by:

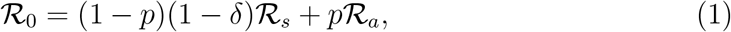

where *ℛ*_*s*_ = *β*_*s*_*/γ*_*s*_ and *ℛ*_*a*_ = *β*_*a*_*/γ*_*a*_ represent the reproduction numbers of asymptomatic and symptomatic individuals (i.e., the average number of secondary infections caused by asymptomatic and symptomatic individuals); therefore, in the absence of the behavioral effect (*δ* = 0), the final size decreases with the asymptomatic proportion *p* because more symptomatic infections leads to a higher basic reproduction number. This relationship changes as *δ* increases. In particular, when *δ >* 1 − *ℛ*_*a*_*/ℛ*_*s*_ (in this case, *δ >* 0.25), the basic reproduction number (and thus epidemic size) increases with *p* because the effective symptomatic reproductive number (including behavioral response) is less than that the asymptomatic reproductive number. For high values of *δ*, we can find a critical level of asymptomatic proportion, *p*_*c*_:

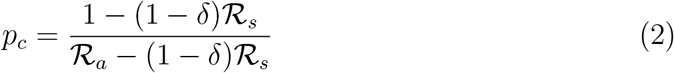

such that *p > p*_*c*_ is required for an outbreak (see threshold effects for large values of *δ* in Fig. 2B).

When behavioral protection is high, the effect of asymptomatic proportion on fatalities shows countervailing effects of individual-level protection and population-level risk (Fig. 2C). For high values of *δ*, the peak fatality occurs at intermediate levels of asymptomatic spread: although fewer individuals die per infection for higher values of *p*, the increase in total infections also leads to an increase in total fatalities. In contrast, when *δ* is small enough such that (1 − *δ*) *ℛ*_*s*_ *≥ ℛ*_*a*_ (in this case, *δ <* 0.25), then total fatalities decrease with *p* because because both the number of infections and the IFR ((1 − *p*)*f*) decrease with increasing *p*.

High values of *δ* required for the nonlinear effects of asymptomaticity on deaths may seem unrealistic. For this particular model, it does not make biological sense for *δ* to be greater than the amount of post-symptomatic transmission, because pre-symptomatic transmission is implicitly included in the *I*_*s*_ compartment. While several studies have estimated the proportion of pre-symptomatic transmission to be around 30%–60% for the SARS-CoV-2 wildtype strain, many of these were likely affected by intervention and behavioral effects, as they were conducted after SARS-CoV-2 awareness became widespread [16]. Instead, [17] recently estimated that the proportion of pre-symptomatic transmission could have been as low as 20% (95%CI: 6%–32%) during the first few weeks of the pandemic when the pandemic-awareness and intervention measures were minimal. There are two implications of this updated estimate—first, a low proportion of pre-symptomatic transmission suggests that high *δ* values are feasible (although not necessarily likely) during the initial pandemic phase; and second, intermediate levels of behavioral effects (*δ >* 0) would have been already present early in the pandemic to reduce the proportion of pre-symptomatic transmission from 80% to as low as 40%.

We therefore extend our model to consider the effects of generalized *non-symptomatic* transmission, which includes both pre-symptomatic and asymptomatic transmission, to ask the following question: does an intermediate amount of non-symptomatic transmission lead to a peak in fatalities? For this model, we assume that *δ* decreases transmission only after symptom onset. We then fix the reproduction number of symptomatic individuals and calculate fatalities at the population level as a function of the proportion of total non-symptomatic transmission and the proportion of non-symptomatic transmission that is caused by pre-symptomatic transmission (see Supplementary Text for model details and Supplementary Table S2 for parameter descriptions and values). Using the generalized non-symptomatic transmission model, we find a wide variety of scenarios for which peak fatalities occur at intermediate levels of non-symptomatic transmission in the presence of moderate to strong behavioral effects, *δ >* 0.6 (Supplementary Figure S1). One exception is the extreme (and unrealistic) case, in which all non-symptomatic transmission is caused by pre-symptomatic transmission (i.e., there are no asymptomatic cases); in this case, total infections and fatalities are maximized when all transmission is caused by pre-symptomatic transmission. Hereafter, we focus on asymptomatic infections for simplicity, but our conclusions have implications for the more general case of non-symptomatic transmission.

We now apply our framework to understand the impact of immunity on total fatalities at the population scale by dividing the population into two groups: immunologically naive and protected. For simplicity, we do not distinguish whether the immunity is derived from natural infections or vaccines. The dynamics of immunologically naive individuals are equivalent to our original model (Fig. 2). The dynamics of protected individuals include three additional parameters, which characterize the amount of protection against infection *E*_*i*_, symptoms (given infection) *E*_*s*_, and deaths (given symptoms) *E*_*d*_ (Fig. 3). For simplicity, we assume that the population is split in half (50% naive and 50% protected) and mixes homogeneously. We also do not consider the separate effect of immunity on transmission (beyond the effect on infection). In other words, we assume that asymptomatic infections in protected and unprotected people have the same reproduction numbers (and likewise for the symptomatic infections). In practice, both asymptomatic and symptomatic infections in protected people are less likely to transmit than their unprotected counterparts [18]: asymptomatic infections in protected people may indicate limited viral replication or even immune boosting, in which case an exposed individual may successfully fight off the pathogen early in infection before it can be transmitted; and symptomatic infections in protected people may reflect a strong immune response (rather than high viral load), in which case symptomaticity can be a poor proxy for transmission. We assume a relatively strong behavioral effect *δ* = 0.8 for illustration.

**Figure 3:**
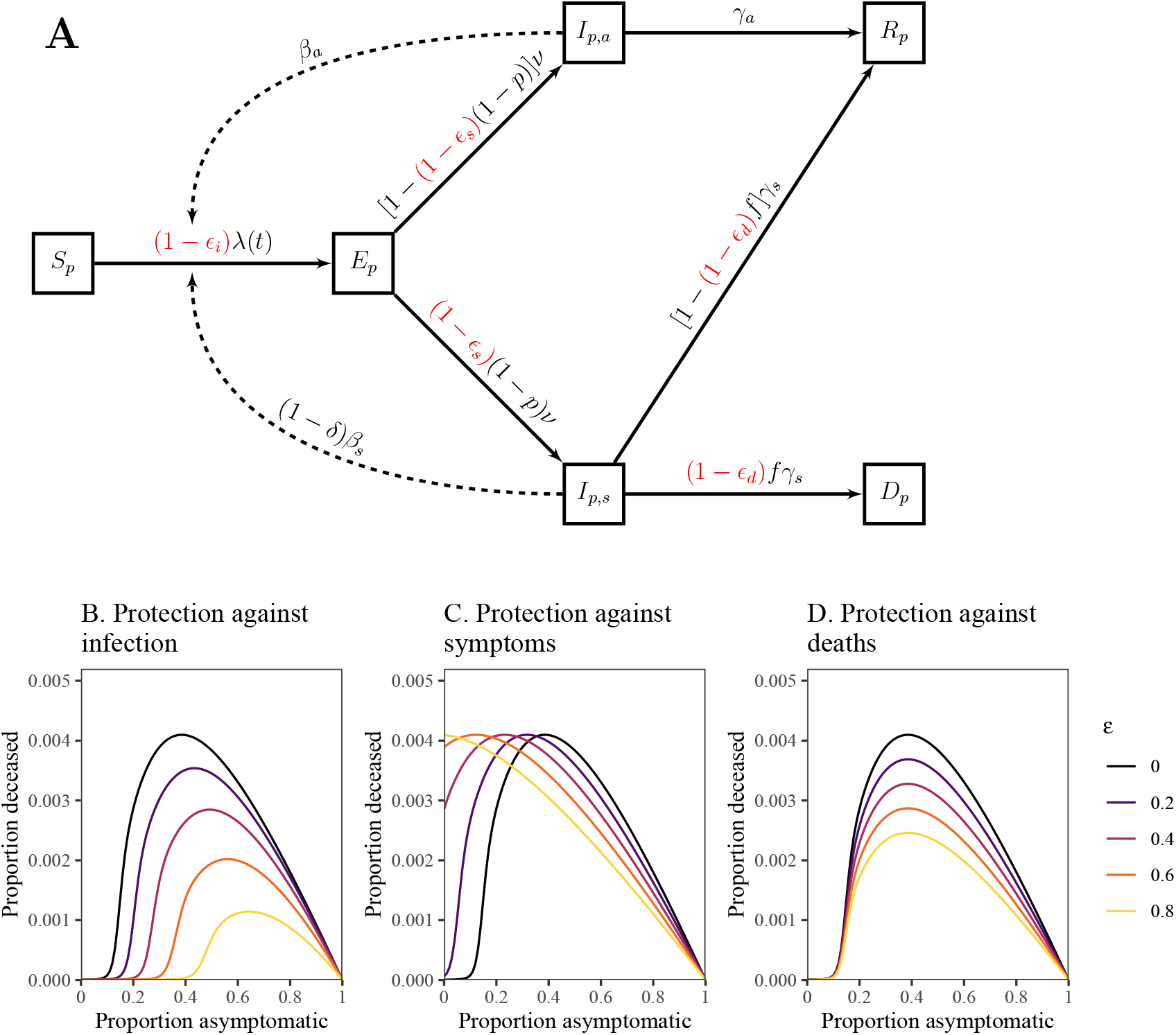
Schematic diagram and simulations of a model with symptom-responsive transmission reduction and immunity. (A) The subscript *p* represents protected individuals. Immunity may provide protection against infection, symptoms, or deaths. The dynamics of immunologically naive individuals are described in Fig. 2. (B–D) Total deaths as a function of the proportion of asymptomatic infections *p* across a wide range scenarios for protection against infection *E*_*i*_ (B), symptoms *E*_*s*_ (C), and deaths *E*_*d*_ (D). We simulate the model for 365 days, assuming *β*_*s*_ = 4*/*5*/*day, *β*_*a*_ = 0.75*β*_*s*_, *ν* = 1*/*2*/*day, *γ*_*s*_ = *γ*_*a*_ = 1*/*5*/*day, *f* = 0.01, and *δ* = 0.8. We assume that 10^−4^ proportion of individuals are initially infected. See Supplementary Text for model details and Supplementary Table S3 for parameter descriptions and values.

We consider each protection effect—*E*_*i*_, *E*_*s*_, and *E*_*d*_—separately and consider joint effects later on. The impact of protection against infection *E*_*i*_ is analogous to changing *ℛ*_0_ in the original model: as immunity provides stronger protection against infection (higher *E*_*i*_), the number of deaths decreases and a higher asymptomatic fraction *p* is required for the infection to spread (Fig. 3B). We note that protection against infection scales the fatality curve nonlinearly, reflecting the nonlinear relationship between *ℛ*_0_ and the final size of the outbreak. The impact of protection against symptoms *E*_*s*_ is equivalent to changing the asymptomatic fraction *p* for the protected population because protected individuals are less likely to develop symptoms: the peaks of the fatality curves move to lower values of *p* as we increase the degree of protection *E*_*s*_ (Fig. 3C). Therefore, for low values of *p*, protection against symptoms can increase the total number of fatalities at the population level by increasing the proportion (and number) of asymptomatic individuals, who can readily transmit infections to other individuals. This also means that the critical level of asymptomatic proportion decreases, allowing more dangerous infections (with lower *p*) to invade, which would not have been able to spread in an otherwise immunologically naive population. We note that the equivalence between protection against symptoms *E*_*s*_ and fraction asymptomatic *p* relies on our assumption that immunity does not provide protection against transmission. Protection against deaths *E*_*d*_ directly modulates the fatality rate for symptomatic cases and therefore linearly scales the fatality curves (Fig. 3D).

Finally, we use our framework to understand the impact of behavioral effects on invading variants (Fig. 4). In doing so, we first simulate the dynamics of a wildtype variant for 1 year using our base model using identical parameters as in Fig. 2. We then simulate a new variant invading a partially immune population using our extended model (Fig. 3A), where the immunity is solely derived from natural infections caused by the wildtype variant in the first year. We consider two types of variants (which are simulated separately): one with the same severity *p* (variant 1, orange) and a milder one with higher *p* (variant 2, purple).

**Figure 4:**
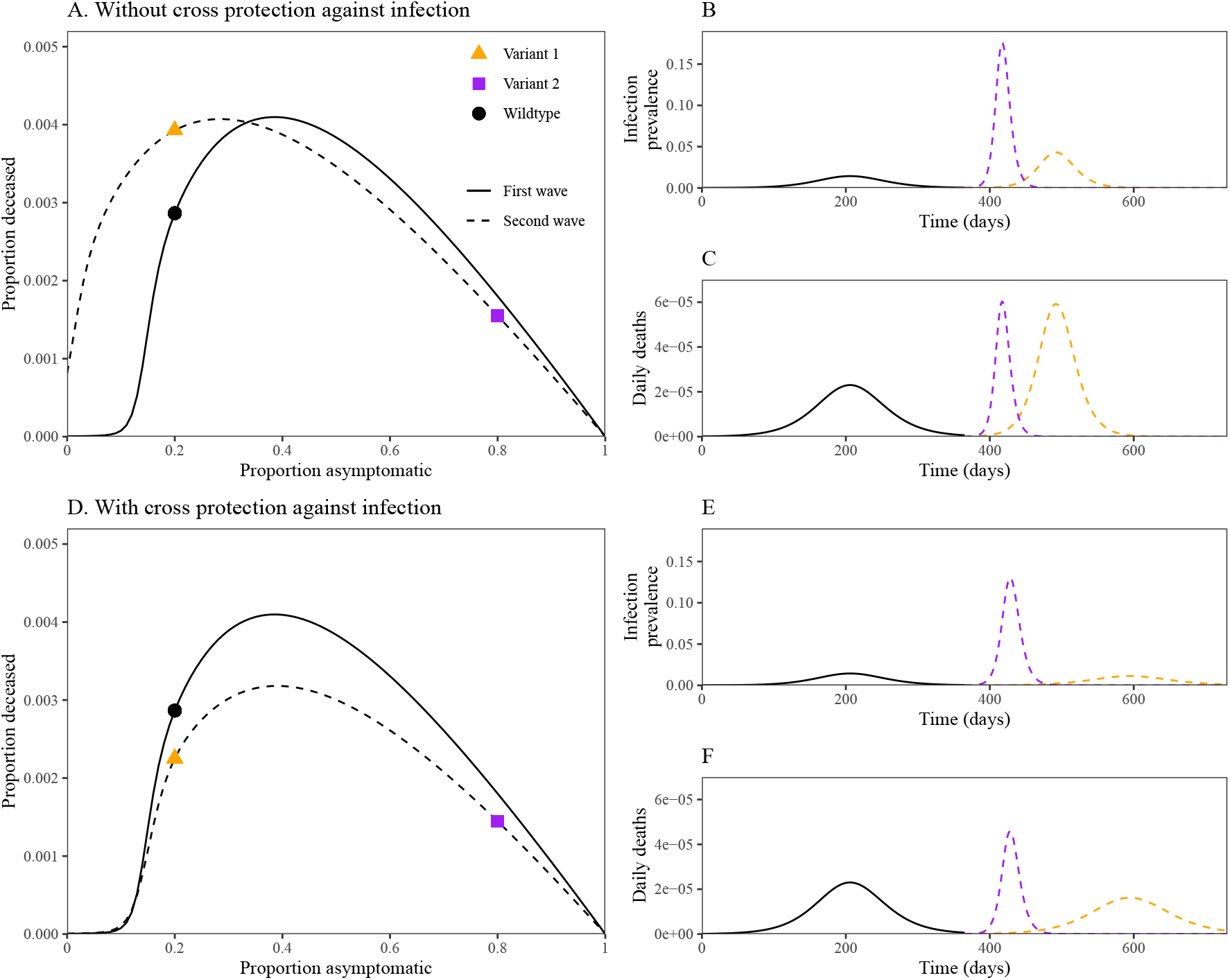
Dynamics of invading variants under symptom-responsive transmission reduction and immunity. (A, D) Asymptomaticity–fatality curves for the first (solid lines) and second waves (dashed lines). Points represent specific scenarios we assume for the first and second waves. Fatality curves for the first wave are calculated by simulating an epidemic for 1 year using parameters from Fig. 2 with *δ* = 0.8. Fatality curves for the second wave are calculated by first simulating the first wave assuming *p* = 0.2 for 1 year to calculate the proportion immune and then simulating the extended model presented in Fig. 3 for two different values of *p* as shown. (B, E) Dynamics of infection prevalence for the wildtype variant (black, solid line) and two possible invading variants (colored, dashed line). (C, F) Dynamics of daily deaths for the wildtype variant (black, solid line) and two possible invading variants (colored, dashed line).

First, we consider a scenario in which immunity only provides protection against symptoms, *E*_*s*_ = 0.4 (Fig. 4A–C). In this case, protection against symptoms allows new variants to spread faster by increasing the amount of asymptomatic infections, resulting in larger outbreaks (Fig. 4B). Although the milder (purple) variant exhibits a faster epidemic growth rate and reaches a higher peak (Fig. 4B), it reaches similar peak fatality as the more severe (orange) variant (Fig. 4C). The asymptomaticity– fatality curve provides additional insight (Fig. 4A): even though a milder, invading variant (purple square) gives higher peak fatality than the original, wildtype variant (black circle), it leads to lower fatalities overall because deaths are concentrated over a shorter period of time in the epidemic. In general, when *δ* is large, invading variants with similar asymptomaticity *p* will spread more effectively and result in worse population-level outcomes if immunity (either from vaccination or natural infection) provides protection against symptoms but not against infection or transmission.

Next, we consider a more realistic scenario in which immunity provides protection against both symptoms, *E*_*s*_ = 0.4, and infection, *E*_*i*_ = 0.4 (Fig. 4D–F). In this case, cross-protection against infection has a large effect on the more severe (orange) variant, causing its peak infection prevalence (Fig. 4E) and fatality (Fig. 4F) to be lower than that of the original, wildtype variant. Across a wide range of asymptomatic proportion *p*, we find that this immunity profile is sufficient to prevent worse outcomes at the population level; we note that the second wave of deaths is still high (and having higher peaks in some cases) even if the overall deaths are lower.

The outcomes in our simulations of invading variants resemble the dynamics of the SARS-CoV-2 Omicron variant. Despite moderate levels of vaccine effectiveness against symptomatic and reduced levels of severe cases caused by the Omicron variant, especially after booster shots [19], both vaccine- and infection-derived immunity provided limited protection against infections [20]. This immune evasion helped the Omicron variant to cause more infections in South Africa than previous variants [21]. Moreover, even though the Omicron variant is probably milder than the Delta variant [22, 23], the number of hospitalizations and deaths caused by the Omicron variant was higher than those caused by the Delta variant in many locations [24, 25, 26].

There are several limitations to our analysis. First, while we are able to generalize the model to include both pre-symptomatic and asymptomatic transmission, behavioral and intervention effects must be relatively large in order for the fatality to peak at intermediate levels of asymptomaticity (typically requiring a reduction in transmission rate of 60% or more for most of our chosen parameter sets). Second, the model framework is able to incorporate the impacts of immunity of infection, symptoms, and severity, but we neglected the effects of immunity on transmission, which also has important effects on disease dynamics [27]. In particular, if immunity provides stronger protection against transmission among immune individuals, population-level outcomes will be better than what our model predicts. Estimating protection against different endpoints (e.g., infection, symptom, death, and transmission) can help narrow this uncertainty. Finally, we assumed that asymptomatic and symptomatic individuals are infected for the same amount of time. Analysis of viral load trajectories suggests that asymptomatic individuals may clear infections faster [7]; however, asymptomatic individuals may still transmit for a longer period of time if symptomatic individuals self-isolate quickly after symptom onset. The individual-level differences in the asymptomatic and symptomatic transmission time scale can have important implications for the inferences and predictions of pathogen dynamics [28, 29]; nonetheless, we expect that predictions on the final size of the epidemic and total fatalities will be robust to small differences in the transmission time scale between asymptomatic and symptomatic individuals.

Even though we assumed a homogeneous population throughout, our analysis also has important implications for age-dependent heterogeneity in asymptomaticity (as shown in Fig. 1D). For example, vaccinations and intervention measures primarily targeting older individuals can prevent severe infections and improve individual-level outcomes. However, asymptomatic individuals, especially younger individuals with high contact rates, can still transmit to other, older individuals, potentially making population-level outcomes worse than they would be if intervention measures were distributed differently. We note that other factors, such as the efficacy of a vaccine and types of immunity provided by the vaccine, also play critical roles in making these decisions—in many cases, protecting the most vulnerable will be the optimal decision to minimize deaths [30].

In summary, using a series of simplified models, we have shown that asymptomatic infections (or, more generally, non-symptomatic transmission) can represent a double-edged sword leading to a better outcome for many individuals while facilitating onward transmission that leads to a worse outcome for the population as a whole. Extending our framework further shows that immunity profile (i.e., reduction of infection, symptoms, and/or severity due to immunity) plays a critical role in determining the dynamics of future variants. For example, while protection against symptoms unaccompanied by protection against transmission protects health at the individual level, it can lead to more infections, and potentially more deaths, at the population level. A similar concern was raised in prioritizing vaccine choices that could reduce severe outcomes vs. others that could reduce transmission [31].

SARS-CoV-2 has proven hard to control in large part because transmission is often decoupled from symptoms. Our model reinforces the need for dual approaches— prioritizing the reduction of asymptomatic spread (e.g., via risk awareness campaigns [32, 33], asymptomatic testing programs [34, 35], mask-wearing indoors and in crowded environments [36, 37, 38], and through improvements in ventilation [39, 40]) while improving the treatment of symptomatic cases, particularly amongst older individuals at highest risk for severe outcomes. Given the link between age and asymptomatic infections [8], interventions may consider different approaches in strongly age-structured populations (e.g., schools or long-term care facilities). Mass vaccination is also expected to be important especially if future vaccines induce more transmission blocking. As more variants continue to emerge, monitoring the impacts of preexisting immunity (whether through vaccination and/or infections) on preventing infections, and not just diseases, will be critical to controlling the course of the pandemic [41].

## Data Availability

All data and code are stored in a publicly available GitHub repository (https://github.com/parksw3/asymptomaticvariant).

## Acknowledgements

JSW acknowledges support from the Chaire Blaise Pascal Program of the Île-deFrance region. The funders had no role in study design, data collection and analysis, decision to publish, or preparation of the manuscript.

## Supplementary Text

### Epidemic models with asymptomatic infection and transmission in the absence of immunity

First, we consider a compartmental model with asymptomatic and symptomatic infections in a homogeneously mixing population. The model dynamics are as follows:

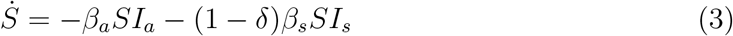

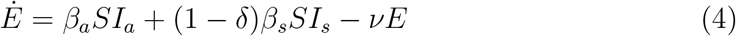

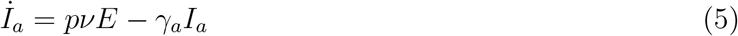

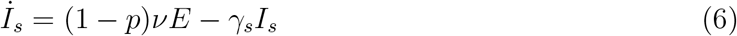

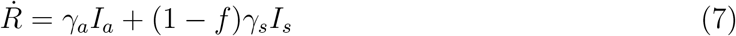

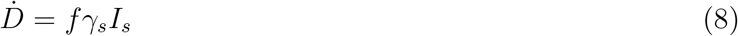

where the transmission rate *β* and removal rate *γ* can be potentially differ between asymptomatic and symptomatic individuals. Here, *δ* denotes the reduction in transmissibility due to responsive measures taken by symptomatic individuals. Throughout the paper, we use parameters that are broadly consistent with the dynamics of the originating strain of SARS-CoV-2: *β*_*s*_ = 0.8*/*day, *β*_*a*_ = 0.75*β*_*s*_, 1*/ν* = 2 days, 1*/γ*_*s*_ = 1*/γ*_*a*_ = 5 days, and *f* = 0.01 [42]. Under this parameterization, we have symptomatic and asymptomatic reproduction numbers of *ℛ*_*s*_ = 4 and *ℛ*_*a*_ = 3.

We then extend the first model to include both pre-symptomatic and asymptomatic transmission:

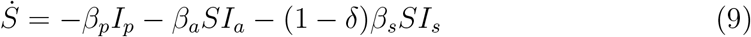

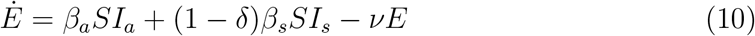

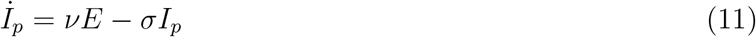

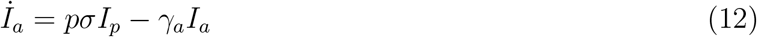

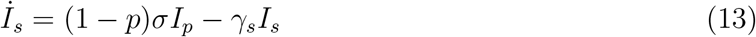

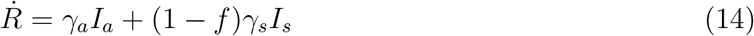

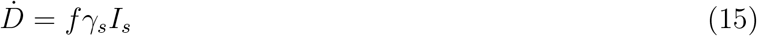

For this model, the pre-symptomatic *ℛ*_*p*_, symptomatic *ℛ*_*s*_, and asymptomatic *ℛ*_*a*_ reproduction numbers are given by _*p*_ = *β*_*p*_*/σ*, _*s*_ = *β*_*s*_*/γ*_*s*_, and _*a*_ = *β*_*a*_*/γ*_*a*_ in the absence of the behavioral effect; these reproduction numbers represent the average number of secondary cases caused by an infected individual in each compartment. Then, the reproduction number of individuals who will eventually develop symptoms is equal to: *ℛ*_*p*_ + *ℛ*_*s*_; similarly, the reproduction number of individuals who remain asymptomatic is equal to: *ℛ*_*p*_ + *ℛ*_*a*_. Since the proportion *p* of all infections is asymptomatic, the basic reproduction number is given by the weighted average of these two reproduction numbers:

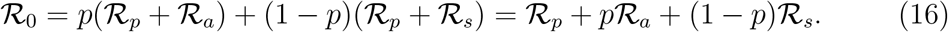

Then, the proportion of non-symptomatic transmission *ϕ* is given by:

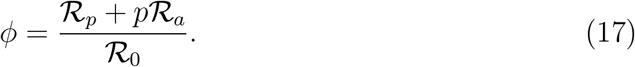

For simulations of the combined model, we start by fixing the reproduction number of individuals who will eventually develop symptoms: _symp_ = _*p*_ + _*s*_ = 4. Consistent with previous assumptions, we also assume that asymptomatic reproduction number is lower than that of the symptomatic reproduction number: _*a*_ = *ρ* _*s*_ where *ρ* = 0.75. Then, for a given value of the proportion of non-symptomatic transmission *ϕ* and proportion of non-symptomatic transmission caused by the pre-symptomatic transmission, *η* = _*p*_*/*(_*p*_ + *p* _*a*_), we can solve for the transmission rate for each compartment *β* and the proportion asymptomatic *p*. More specifically:

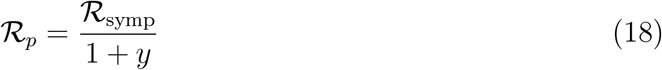

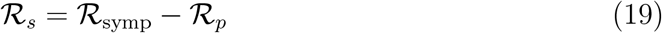

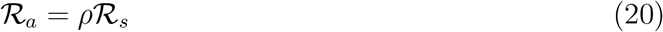

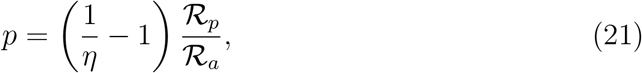

where *y* = (1*/ϕ* − 1)*/η* + (1*/η* − 1)*/ρ*. In order to keep the mean infectious period fixed, we assume 1*/σ* = 2 days and 1*/γ*_*s*_ = 1*/γ*_*a*_ = 3 days. All other parameters are same as before.

### Epidemic models with asymptomatic infection and transmission in the presence of immunity

We then model the spread of infection in a partially immune population. The model dynamics are as follows:

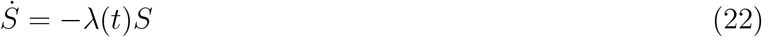

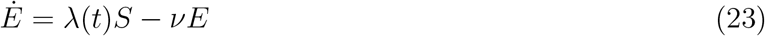

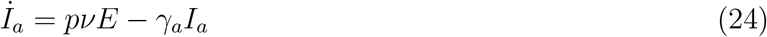

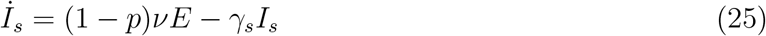

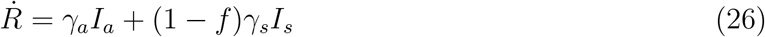

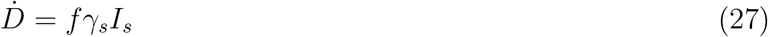

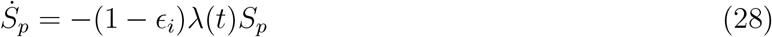

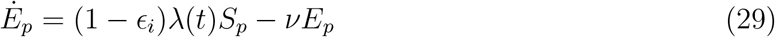

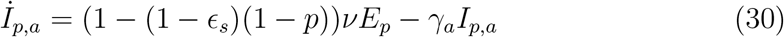

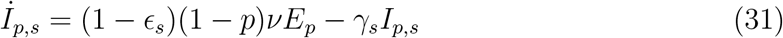

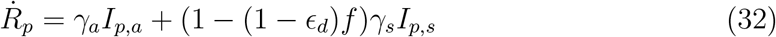

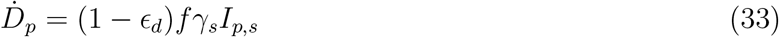

where 0 ≤ *E* ≤ 1 represents the degree of protection against infection, symptoms and death. The force of infection *λ*(*t*) is given by:

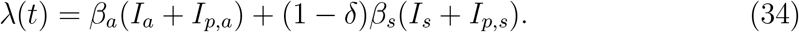

Here, subscripts *p* denote individuals who are immune and therefore are protected.

## Supplementary Tables

**Table S1:**
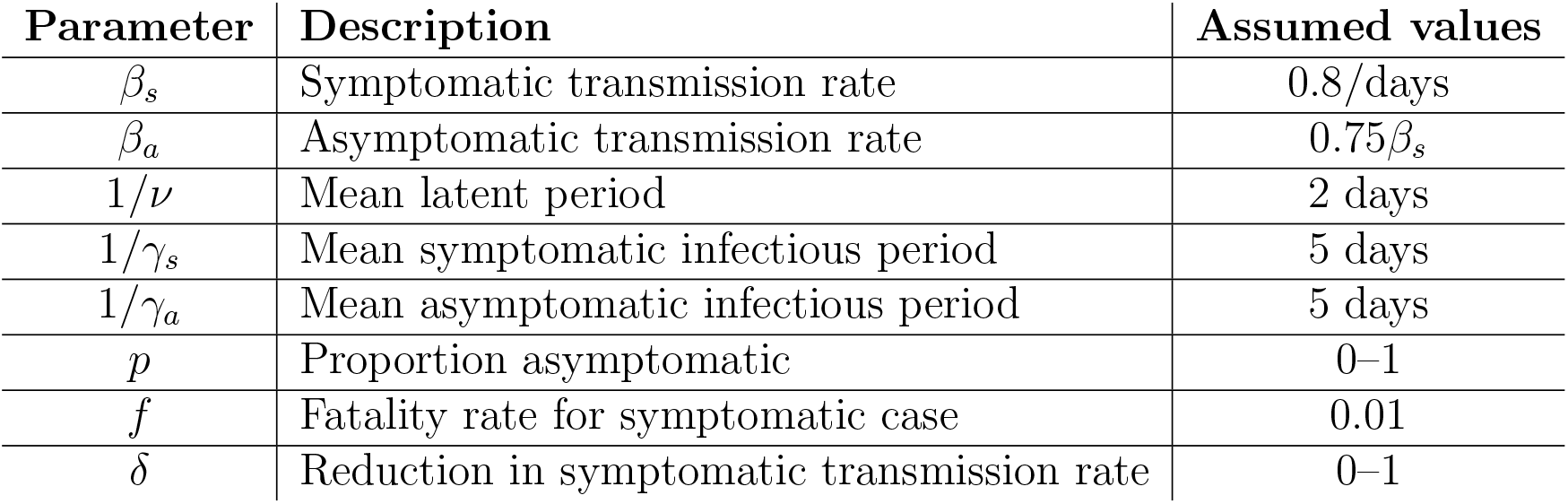
Paramter descriptions and values for the basic asymptomatic model.

**Table S2:** Paramter descriptions and values for the generalized asymptomatic model.

| Parameter | Description | Assumed values |
| --- | --- | --- |
| $\beta_s$ | Symptomatic transmission rate | See Supplementary Text |
| $\beta_a$ | Asymptomatic transmission rate | See Supplementary Text |
| $\beta_p$ | Presymptomatic transmission rate | See Supplementary Text |
| $1/\nu$ | Mean latent period | 2 days |
| $1/\sigma$ | Mean presymptomatic infectious period | 2 days |
| $1/\gamma_s$ | Mean symptomatic infectious period | 3 days |
| $1/\gamma_a$ | Mean asymptomatic infectious period | 3 days |
| $p$ | Proportion asymptomatic | 0–1 |
| $f$ | Fatality rate for symptomatic case | 0.01 |
| $\delta$ | Reduction in symptomatic transmission rate | 0–1 |

**Table S3:**
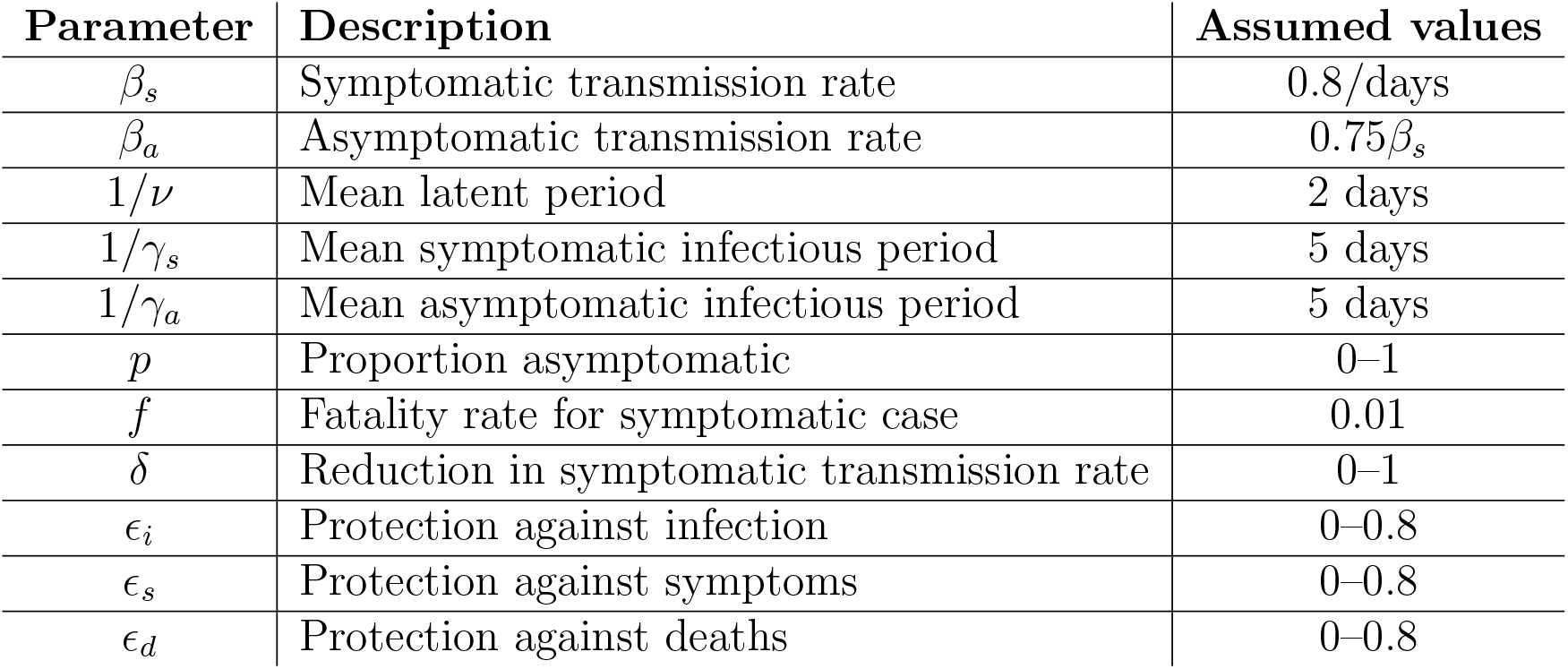
Paramter descriptions and values for the asymptomatic model with immunity.

## Supplementary Figures

**Figure S1:**
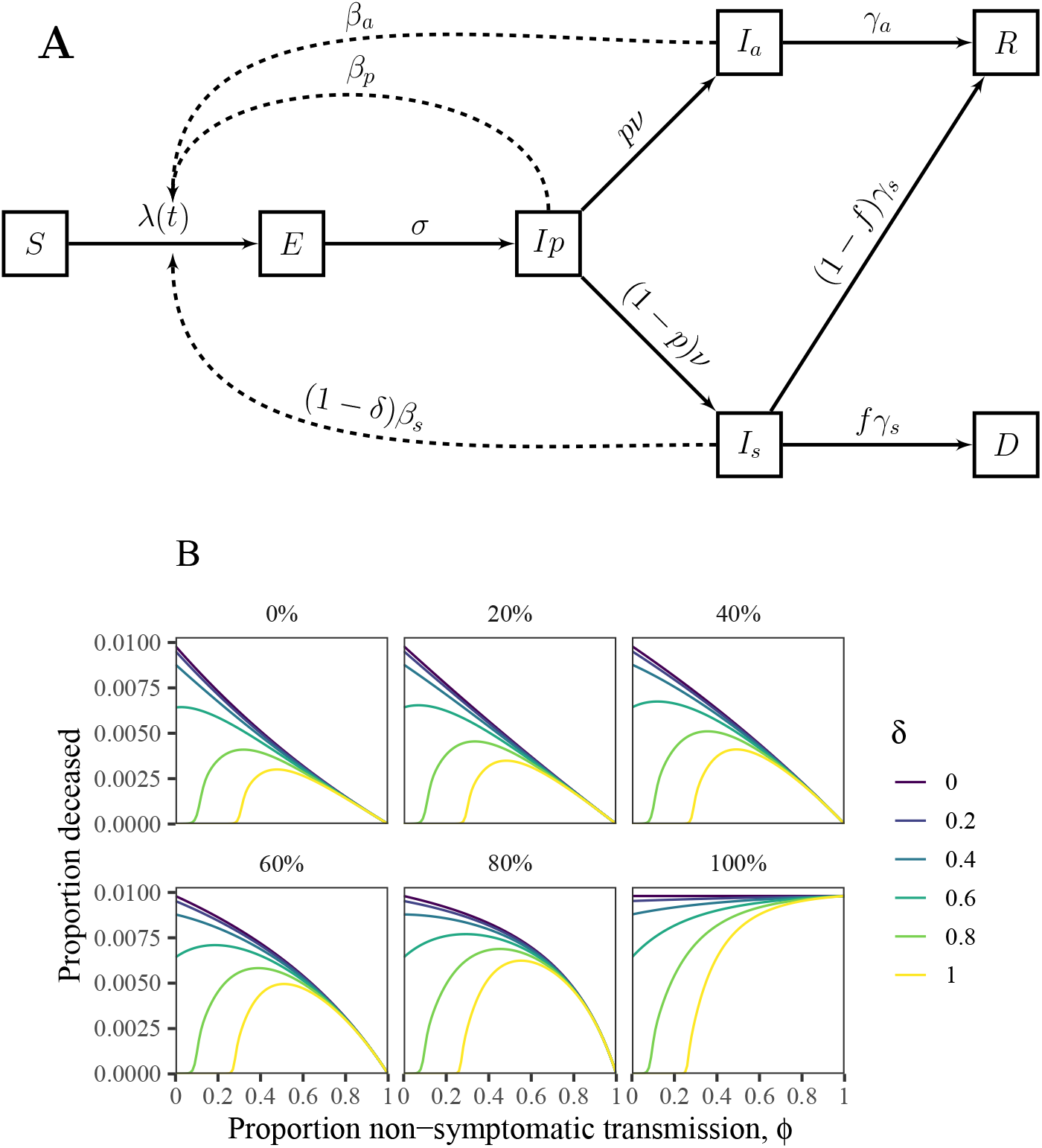
Schematic diagram and simulations of a model with pre-symptomatic and asymptomatic transmission and symptom-responsive transmission reduction. (A) *S* represents susceptible individuals; *E* represents exposed individuals; *I*_*p*_ represents pre-symptomatic individuals; *I*_*a*_ represents symptomatic individuals; *I*_*s*_ represents symptomatic individuals; *R* represents recovered individuals; and *D* represents deceased individuals. See Methods for model details. (B) Total deaths as a function of the proportion of non-symptomatic transmission *ϕ* across a wide range scenarios for *δ* and proportion of non-symptomatic transmission caused by the pre-symptomatic transmission, *η* (between 0% and 100%). See Supplementary Text for model details and Supplementary Table S2 for parameter descriptions and values.

